# Assessment of knowledge, attitude, and practices of Tuberculosis patients towards DOTs regimen in Jimma Health Center, Jimma zone, southwest Ethiopia

**DOI:** 10.1101/2021.07.26.21261113

**Authors:** Wudalem Amare, Melese Sinaga Teshome, Gashaw Dessie, Tabarak Malik

**Affiliations:** Department of Population and Family Health, Nutrition Unit, Jimma University, Jimma, Ethiopia; Department of Biochemistry, School of Medicine, College of Medicine and Health Sciences, University of Gondar, Gondar, Ethiopia

**Keywords:** Knowledge, Attitude, Practice, TB, Dots regimen

## Abstract

**Objective:** Globally, tuberculosis (TB) is one of a chronic infectious disease, which has a major health burden. Thus, 9.4 million incidents and 14 million prevalent cases were reported in 2010. The high burden of TB is common in sub-Saharan Africa countries, including Ethiopia. The purpose of conducting this study was to assess the knowledge, attitude, and practices of TB patients towards DOTs regimen. The other objective of the study was to assess the attitude of patients towards causative factor and its treatment.

**Results:** The result of the study showed that 103 (68.7%) of patients scored high knowledge status, whereas 47 (31.3%) of them scored low. 25(16.7%) governmental employee had overall high knowledge status. 101 (67.3%) of the patient scored high overall attitude, while 49 (32.7%) of them scored low overall attitude. 85 (56.7%) of patients scored high overall practices, whereas 65(43.3%) of them scored low overall practices.In this study, the knowledge and attitude of patients about TB and its treatment was found within low level of perception. Therefore, implementation of health education and awareness creation are required to avoid the problem of patients.

## Introduction

Tuberculosis (TB) is a chronic infectious disease caused by *Mycobacterium tuberculosis*, whichleads tohigh public health burden in developing countries. The incidence of TB cases expected to be reached around one billion from 2002 to 2020 (1). Globally, one third of the world’s population is expected to be infected with tubercle bacilli, thusthe risk of developing active TB is high. Annually, 8.4 million people develop active TB and 2.3 million died from the disease. TB comprises 25% of adult’s death in developing countries and it is the leading cause of death among young women. Its burden is highly elevated particularly among TB/HIV co-infected young women (2). Immune-competency and proper nutritional diet is required for prevention and treatment of TB (3).TB affects individuals atall age group, sex andall socio-economic status. It is the most common cause of death in an individualwithin15-54 years old, which may be leads to high socio-economic consequences. Studies conducted in Gambella showed that the prevalence of pulmonary TB cases were higher within 15-24 and 25-34 years old (4).

According to the World Health Organization, the incidence of TB was estimated to be 10.4 million cases and 1.4 million populations had been died in 2015respectively(5). World Health Organization has declared TB as a global public health emergency to control the elevation of TB cases, thereforethe organizationdesign a new strategic framework for effective control and management(6). According to WHO, over 30 million patients are treated usingDirectly Observed Treatment programme. In this programme, the default and curative rates of TB are expected to be <10% and >80% respectively(7). It is a vital for successful TB control to treatit within this framework in national TB program(8).

Drugs utilized for the treatment of DOTS system are not novel drugs. They have been available for last the three decades, and theyhave been almost 100% effective.The proper use of these drugs often limited in hospitalsbecause many challengescreate burden to carefully monitored regimens and patient progress. Monitoringof patients in the hospital for two months leads to overcrowdon the TB ward, and it may alsoinfluenceon the patient’s family, social and economic situation. DOTS require less time in the hospital for supervised treatment(8).At the beginning of programme (1993) at a global level, more than 2% of the active TB cases were not treated by this method. According to World Health Forum reported in 1997, the estimated figure is nearly 12%, which reflects a remarkable rate of expansion. Millions of TB patients still require the accessibility of the system(9). Therefore, this study was conducted in Jimmahealth centerto assess the knowledge, attitude, and practices of TB patients towards DOTS regimens.

## Methods and materials

### Objectives of study

The purpose of conducting this study was to assess the knowledge, attitude, and practices of TB patients towards DOTs regimen. The other objective of the study was to assess the attitude of patients towards causative factor andits treatment.

### Study design

A cross-sectional study was conducted based on the patient card and registration at TB follow-up clinic. It was conducted from June 20 to September 19, 2016.

### Study population

All TB patients who registered on DOTS regimen and had known treatment outcome at the Jimma health center TB follow-up clinic were included duringthe study period.

### Eligibility criteria for study participants

#### Inclusion criteria

All registered TB patients who were volunteer and started treatmentinvolved in this survey.

#### Exclusion criteria

Exclusion of study participants was done carefully by professionals.Seriously ill-patientswere excluded in this study.

### Data collection procedure and process

In this study, well-structuredquestionnaireformat was prepared. It was composedof socio-demographic characteristic, treatment outcome, and category of the patient. All TB patients registered at the TBfollow-up clinic were identified from the registration book.

### Data Processing and Analysis

Data was processed and analyzed manually. In addition to this, odd ratio (OR) with 95% CI was computed to assess the degree of association.

### Ethics approval and consent to participate

Ethical approval was obtained from the research and ethics committee of the School of Medicine, Jimma University, Ethiopia. Consent forms are available from corresponding author on reasonable request and all participants were provided written informed consent. Department of Ethics and Research Committee decided and approve the protocol.

## Results

### Socio-demographic characteristics

In this study, a 100% response rate was detected from thetotal 150 respondents, 70 (46.7%) werewithin age interval of 15-45. From the total enrolled study participants, 70 (46.7%) were within 15-49, and 54 (36%) of them were above 50 years old, and 26 (17.3%) of them were less than 14 years old. Regardingreligion and ethnicity, 49 (32.7%) were Muslim, and 72 (48%) were Oromo. Out of 150 study subjects, 89(59.3%) are married and 142(94.7%) were able to read and write. On the other hand, 37 (24.7%) were housewives, only 6(9%) of them had <150 birr monthly income, whereas 10(6.6 %) of them had>1500.

### Distribution of DOTS regimen by sex and types of diagnosis

Out of the total of 76 study subjects, PTB positivity was identifiedbydiagnosis type and sex.The results showed that 45 (30%) males and 31(20.7%) females were positive for TB, whereas 15 (10%)males and 25 (16.7%) females were negative. 19 (12.7%) males and 15 (10%) females were extrapulmonary (EP) cases.

### Comparision of age group of TB patient and treatment outcomes

Regarding evaluation of the treatment outcome, the higher cure rate was observed between 15-49 years old. On the other hand, the treatment completion rate was higher in >50 years old.Thedeath rate was higheratthe old age groupcatagory.

### Comparision of treatment outcomes with types of TB diagnosis

Out of the total 76 pulmonary positive TB (p/Pos)patients, 43 (28.7%) were cured and 2 (1.3%) of them were died.Similarly, out of 40 pulmonary negative TB patients (p/Neg),22 (14.7%) were under cured treatment course. From the total 34 EP cases, 27 (18%)were completedthe treatment course successfully.

### Patient’s overall knowledge and attitude about Tuberculosis and its treatment

The result of thisstudy showed that122 (81.3) of patients reported that the transmission of tuberculosis is through bacilli. Of total study participants, 129 (86%) and 143 (95.3%) patients consideredTBas a curable disease, and life treating respectively. Similarly, 148 (98.7%) patients reported duration of treatment requires 6-months, whereas120 (80) patients respond treatment duration takes>6 months. In this study, 103 (68.7%) patients scored above the mean knowledge status, whereas 47 (31.3%) patients had below the mean. Similarly, patient’s overall attitudewas categorized as a high attitude status.

## Discussion

According toWHOreport, women in reproductive age group have a higher risk of developing TB than men within the same age group.However, the incidence of TB in China showedthat men is 27% more likely to develop the disease than women within 25-44 years old (10).It may be associated with HIV/AIDS-related factor because males aremore risky and lower detection rate of disease. From the total 70 TB patients,37 (46.8%) of them were males, whereas 33 (46.5%)of them were females. Additionally, 70 (46.7%) study subjects were from 15 to 49,whereas 54 (36%) of them were above 50 years old.In this study, a diagnostic classification of study subjects were utilized to evaluate TB as a positive and negative results. It was revealed that the highest positive TB cases were found between 15-49 years old. Thus, it was agreed with previous study (11).The result of this study showed that 45 (30%) of males and 31(20.7%) of females were positive for TB, whereas 15(10%) of males and 25(16.7%) of females were negative for TB cases.In addition, 19 (12.7%) of males and 15 (10%) of females were identified as EP cases (figure 1). High pulmonary positive cases had detected among economically active reproductive age group. It may be because ofthose study participants areexposed toHIV/AIDS, which has high degree of co-infection with TB.The study done in China (2017) showed the presence of high prevalence of TB among patients who are co-infected with HIV (12).High proportion of TB cases were identifiedabove 50years old, which accounts up to36% of them.

**Figure 1.**
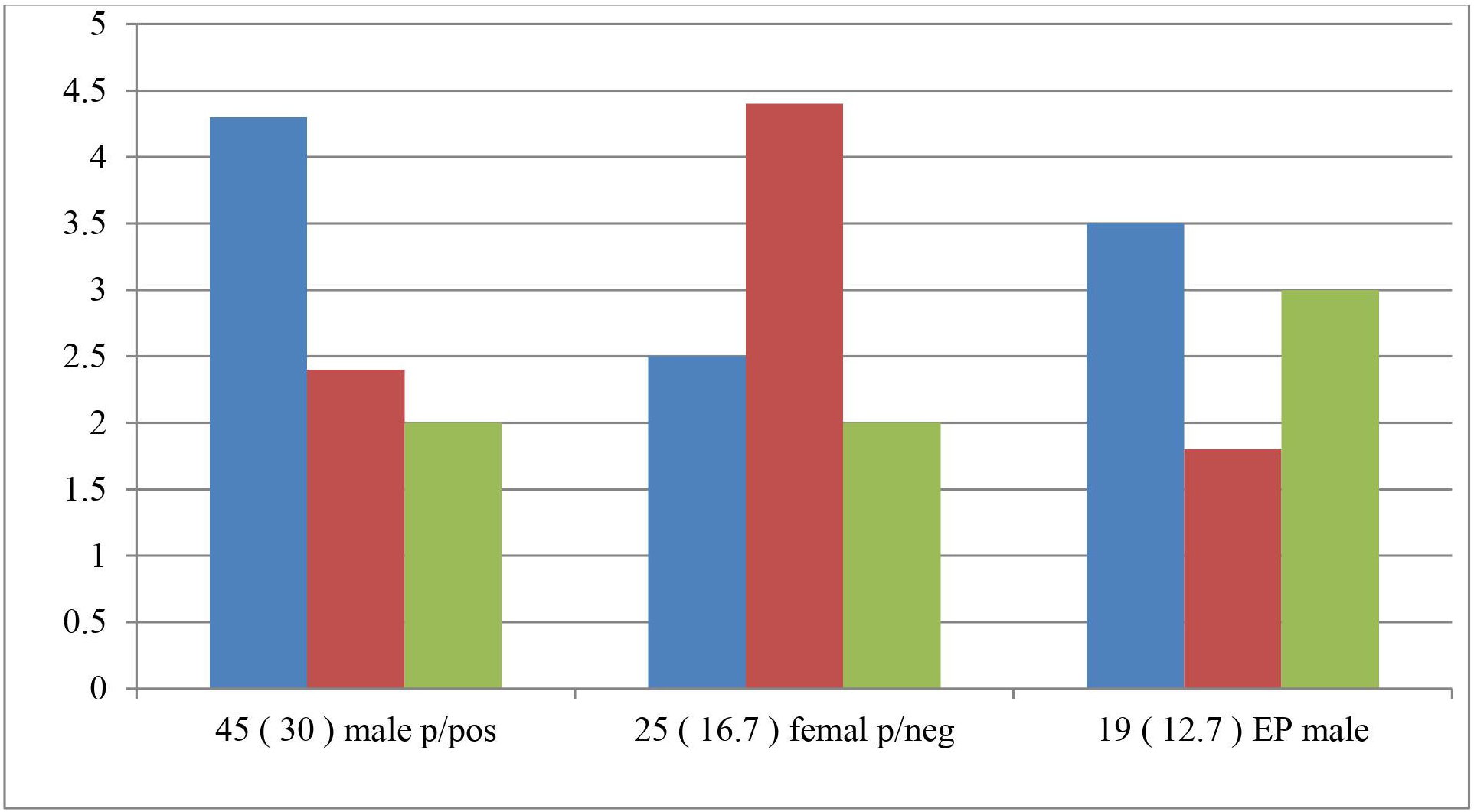
Distribution of patients on DOTS regimen by sex and types of diagnosis.

Studies notified that DOTS increasesthe cure rate of patientsin countries, like Mozambique and Tanzania havinga cure rate of 90% and 80% respectively(13). In this current study, 67(44.7%) and 73(48.7%) study participants were cured and completed their treatment respectively (Table 1).The major challenge of DOTS is the failarityof the program due to inadequite supervision cases until they complete the treatment (14).Out of the total 76 PTB patients (p/pos), 43 (28.7%) were cured and 2 (1.3%) of them died. Out of 40 PTB patients (p/Neg), 22 (14.7%) were cured. Out of total 34 EP cases, 27 (18%) were completed the treatment successfully(Table 2).

**Table 1:** Evaluation of age category according to treatment outcomes

| Age | Treatment outcome |  |  |  |
| --- | --- | --- | --- | --- |
|  | Cured | Completed | Death | Total |
|  | No (%) | No (%) | No (%) | No (%) |
| 0-14 | 9(6) | 15( 10) | 2(1.3) | 26(17.3) |
| 15-49 | 41<br>(27.3) | 26(17.3) | 3(2) | 70(46.7) |
| 50+ | 17<br>(11.3) | 32(21.3) | 5(3.3) | 54(36) |
| Total | 67<br>(44.7) | 73(48.7) | 10(6.7) | 150(100) |

( Descriptive analysis was done to evaluate the outcome of treatment within different age interval)

**Table 2.** Types of diagnosis TB patient on DOTS regimen and treatment outcome

| Types of diagnosis | Treatment outcomes |  |  | Total |
| --- | --- | --- | --- | --- |
|  | Cured | Completed | Death |  |
|  | No (%) | No (%) | No (%) | No (%) |
| p/pos | 43 (28.7) | 31(20.7) | 2(1.3) | 76(50.7) |
| p/Neg | 22 (14.7) | 15(10) | 3(2) | 40(26.7) |
| Ep | 2(1.3) | 27(18) | 5(3.3) | 34(22.6) |
| Total | 67 (44.7) | 73(48.7) | 10(6.6) | 150(100) |

(Descriptive analysis was done. p/pos; Smear positive Pulmonary Tuberculosis, p/Neg; Smear Negative pulmonary Tuberculosis, Ep; Extra Pulmonary )

Theresult of this study showed that overall knowledge of patient about TB was unsatisfactory, but they had better perception than the study conducted on Amhara region in Ethiopia(15). However, they had lower perception than the study conducted in Iraq (16). In this study, 81.3% of patients mentioned bacteria as the cause of TB, whereas 33.7% patients in Addis Ababa explained it as a causative factor(17).In Nigeria, 50% of patients had better knowledge statuson the etiological agent of TB(18). 60% of Nigerian TB patients had good knowledge about the transmission. It may be due tohigh mobilization on health education forgeneral population. In this study,16.7% of government employees had high knowledge status, because they had high educational status.It was in line with a previous work(17).

In this study, 30.3% of patients had unfavorable attitude about TB and its treatment.Similarly, the study done in Iraq revealed that 54.8% of patients had unfavorable attitudes and practices towards TB. In addition,43.4% of patients in Addis Ababahad unfavorable attitudes about TB.The negative attitude of patient towards health care and TB treatment approach was identified as an important attributable factor for defaulting (19).In this regard, a high rate of default (82%) was identified at Addis Ababa TB center. It may be due to inadequate knowledge, low educational level, and negative attitude towards TB center.Out of total study participants,129 (86%) of them agreed on DOTS can cure TB, and theydisagreed on TB treatment by traditional medicine. On the other hand, the study conducted inthe Amhara regional state showed that 95%and 36.9% of patients agreed on both DOTs and traditional medicines respectively (15).In this study, 18 (12%) of patients discontinued DOTS. In line to this, the previous study done in china also showed that 20% of the TB patients interrupted their treatment (20).It wasalso agreed with study conducted in Amhara regional state, Ethiopia (15).

## Conclusion

The result of this study identified that there was a knowledge gap in areas of causative factor, transmission, and symptoms of TB. Asignificant number of patientshad unfavorable attitude towards the disease and itstreatment. Generally, the overall result of the study indicated the presence of was a low level of knowledge, attitude, and practices of patientstowards DOTs.

## Data Availability

Data are available from the corresponding author on a reasonable request.

## Availability of data and materials

Data are available from the corresponding author on a reasonable request.

## Competing interests

The authors declare that there is no any competing interest.

## Funding

Jimma University funded for this project to support graduate students.

## Acknowledgments

We sincerely acknowledge Jimma University, Jimma health center, and support during the research work. We thank all the participants who were enrolled in this study.

## Notes

### Competing Interest Statement

The authors have declared no competing interest.

### Author Declarations

Ethical approval for the study was obtained from Jimma University ethical review committee and official letter was obtain from department of population and family health.

